# Use of supervised machine learning algorithms in predicting postoperative mortality in gastrointestinal and HPB surgeries

**DOI:** 10.1101/2023.07.05.23292033

**Authors:** Bhavin Vasavada

## Abstract

**Aim of the study:** This study aims to evaluate supervised machine learning algorithms in predicting 90 days post-operative mortality in gastrointestinal and HPB surgeries and comparing them with standard logistic regression methods.

**Methods:** We evaluated various supervised machine learning classification algorithms like gradient boosting, K-nearest neighbours, random forest, and support vector machines with standard logistic regression methods. We used accuracy and the Receiver operating curve to compare the methods. 60% of the data were used for training, 20% for validation and 20% for testing. We used JASP 0.16.04 by the University of Amsterdam to run machine learning algorithms and statistical analysis.

**Results:** We used data from 504 patients who have undergone gastrointestinal and hepatopancreatic biliary surgery between April 2016 and March 2023. We analyzed algorithms for predicting 90 days post-operative mortality based on features like Major surgeries, Surgeries for malignancies, age, CDC grade of surgeries, Intraoperative hypotension, Open vs Laparoscopic surgeries, ASA grade, Emergency surgeries, Operative time, Intraoperative blood product used, colorectal surgeries, small intestinal surgeries, HPB surgeries, upper gastrointestinal surgeries and hernia. Test accuracies were 96% for gradient boosting, 90 % for K-nearest neighbours, 96% for the random forest, 94% for support vector and Areas under the ROC curve were 0.802 for gradient boosting, 0.489 for K-nearest neighbours, 0.934 for random forest and 0.5 for support vector algorithms. Accuracy and Area under the ROC curve with standard logistic regression method were 94% and 0.757. Features of importance in decreasing order were ASA, operative times, blood products, small bowel surgeries and Age.

**Conclusion:** Supervised machine learning algorithms particularly gradient boosting and random forest predicted 90 days post-operative mortality more accurately than logistic regression and such models can be part of the preoperative evaluation in gastrointestinal and HPB surgeries.

## Background

In recent times machine learning has been increasingly used in medical research. In recent times machine learning is found superior to standard statistical methods, particularly regarding complex data structure and helps in studying and making predictions even in nonlinear relationships. Machine learning has many algorithms and is particularly classified into supervised and unsupervised machine learning algorithms. Supervised machine learning methods are used to describe prediction tasks because the goal is to forecast/classify a specific outcome of interest. [1].

There is various type of supervised machine learning algorithms and the support vector algorithm has been applied more frequently but in various studies, the random forest algorithm has shown superior accuracy. [2]

Our study aims to evaluate postoperative mortality using supervised machine learning algorithms in our data and compared them with standard logistic regression statistical analysis to evaluate the usefulness of the model and also evaluate various preoperative and intraoperative features of importance in predicting mortality.

## Methods

We used the data of all the patients operated on for gastrointestinal and hepato-pancreaticobiliary surgery in our institute between April 2016 to March 2023. 60% of the data was used for training, 20% for validation and 20% as a test cohort. We kept 90 days mortality as our target variable. We used various pre-operative and intraoperative features like major surgeries, Surgeries for malignancies, age, CDC grade of surgeries, Intraoperative hypotension, Open vs Laparoscopic surgeries, ASA grade, Emergency surgeries, Operative time, Intraoperative blood product used, colorectal surgeries, small intestinal surgeries, HPB surgeries, upper gastrointestinal surgeries and a hernia for creating the model.

### Definitions

#### Mortality

Nine-day mortality was defined as any cause of mortality in the 90-day postoperative period. Ninety-day mortality included all the in-hospital mortalities.

#### Intraoperative Hypotension

Intraoperative hypotension was defined as systolic arterial pressure below 80 mmHg, a decrease in systolic arterial pressure by 20% below baseline, or vasopressor requirement.

#### Centre of Disease Control Grading

We also defined surgical wounds according to the Centre for Disease Control as clean (grade 1), clean-contaminated (grade 2), contaminated (grade 3), and dirty (grade 4).

#### Major and Nonmajor Surgery

We defined surgeries with literature-proven negligible mortality like laparoscopic cholecystectomy, all hernia surgeries, and laparoscopic appendicectomies as nonmajor surgeries and other surgeries as major surgeries. All emergency surgeries except for the abovementioned surgeries were also defined as major surgeries.

#### Supervised Machine learning models

We evaluated various supervised machine learning classification algorithms like gradient boosting, K-nearest neighbours, random forest, and support vector machines with standard logistic regression methods. We used accuracy and the Receiver operating curve to compare the methods. 60% of the data were used for training, 20% for validation and 20% for testing. As the target variable was categorical, we used classical We used JASP 0.16.04 by the University of Amsterdam to run machine learning algorithms and logistic regression analysis was also done using JASP 0.16.04. We also evaluated the confusion matrix, class proportion, evaluation metrics, deviance and out bag improvement plots, relative influences of features and decision boundaries matrix as per the methods applied. We used out-of-box classification error plots and deviance plots to check model accuracy.

## Results

We used data from 504 patients who have undergone gastrointestinal and hepatopancreatic biliary surgery between April 2016 and March 2023. We analyzed algorithms for predicting 90 days post-operative mortality based on features like Major surgeries, Surgeries for malignancies, age, CDC grade of surgeries, Intraoperative hypotension, Open vs Laparoscopic surgeries, ASA grade, Emergency surgeries, Operative time, Intraoperative blood product used, colorectal surgeries, small intestinal surgeries, HPB surgeries, upper gastrointestinal surgeries, and hernia. Test accuracies were 96% for gradient boosting, 90 % for K-nearest neighbours, 96% for the random forest, 94% for support vector and Areas under the ROC curve were 0.857 for gradient boosting, 0.489 for K-nearest neighbours, 0.767 for random forest and 0.5 for support vector algorithms.

As gradient boosting and the random forest were showing the highest accuracies and AUROC (area under ROC curve) we analyzed them in detail.

### Gradient boosting algorithm

As shown in Table 1, 321 patients as training data, 81 for validation and 100 for the test. Validation accuracy was 97.5% and test accuracy was 96%.

#### Boosting Classification

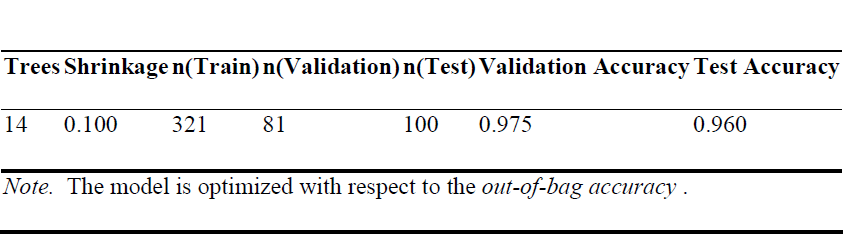

#### Evaluation Metrics

**Table 2.**
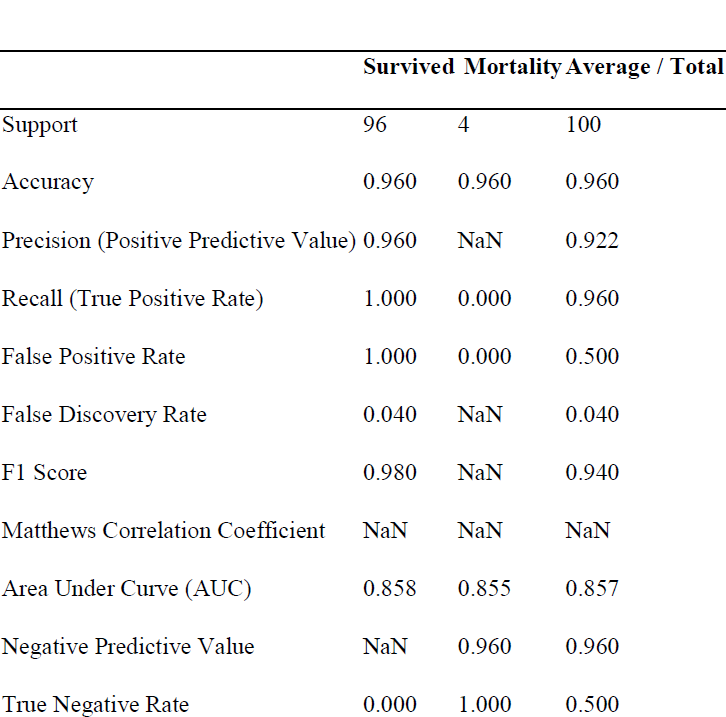
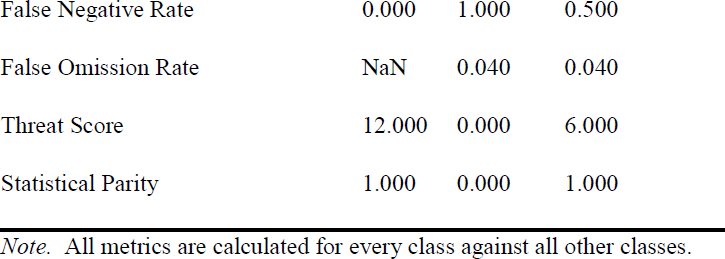
Evaluation Metrices.

As shown in evaluation matrices overall accuracy of the model was 96% precision or the positive predictive value was 92% and the Recall or true positive rate was 96%.

### ROC curve

Now any classification algorithms are biased towards groups with larger sample sizes and hence in the table in the mortality cohort some of the values were not accurately calculated and predicted as the overall 90 days mortality rate in the data was 6.2% so. Naturally, there is a huge gap in numbers in both groups one of the methods to circumvent this problem is to see Area Under Curve in both groups which was highly significant with an overall value of 0.857 and 0.858 and 0.855 in survival and mortality groups.

#### ROC Curves Plot

**Figure 1.**
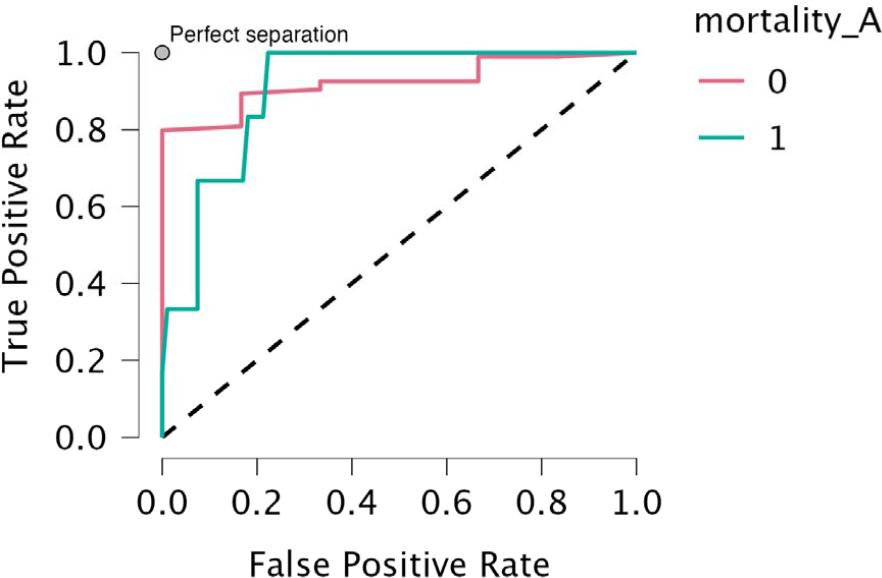
ROC curve of classical gradient boosting.

### Out of box error and deviance plot

Figure 2 shows of box error vs the number of trees plot, as can be seen as the number of trees increased out of box error reduced and was minimal after 14 trees showing the adequacy of the model and adequacy number of trees. The deviance plot also shows a similar trend

#### Out-of-bag Improvement Plot

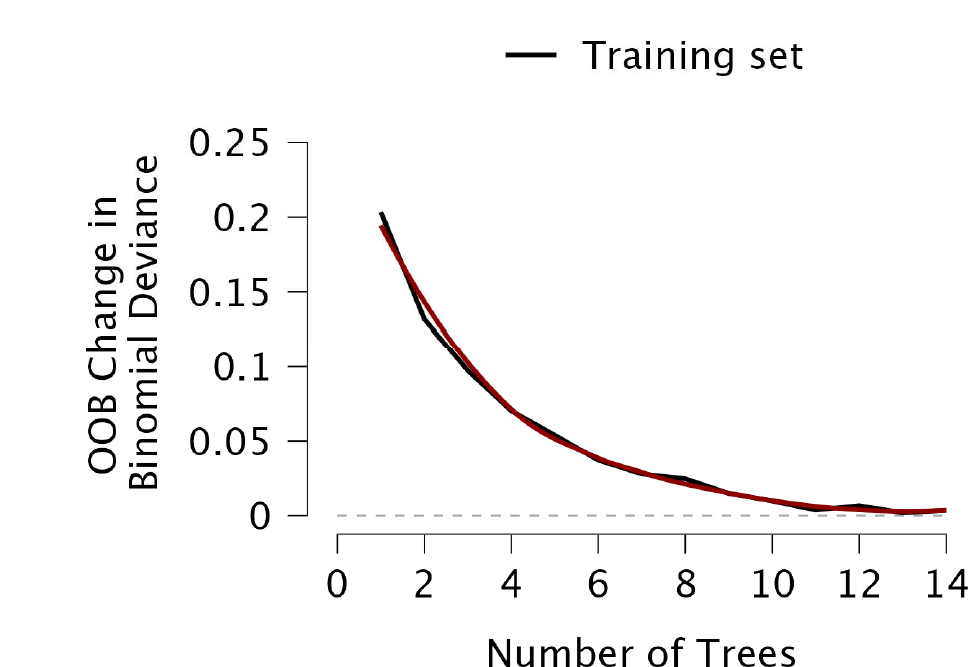

#### Deviance Plot

**Figure 2.**
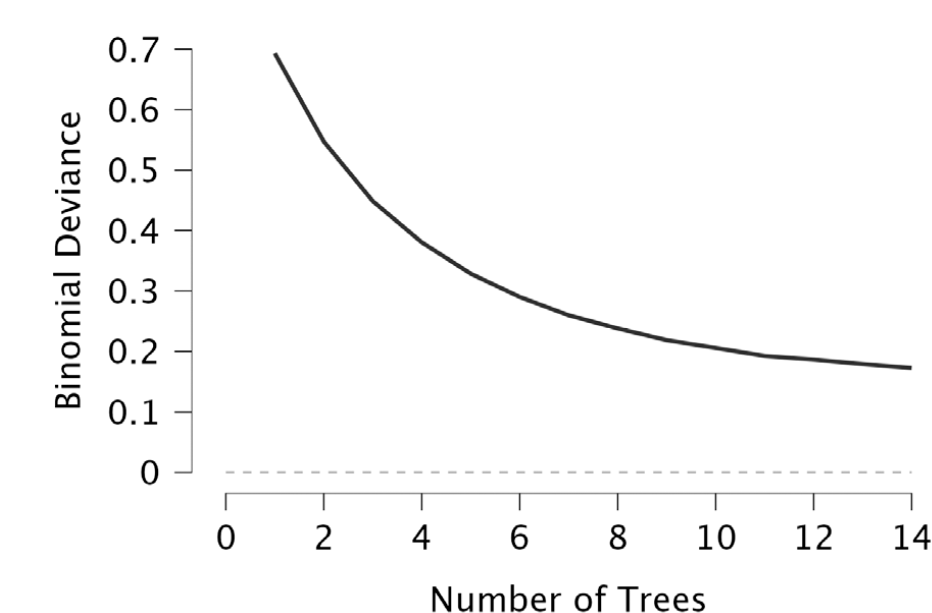
Out of bag improvement plot and deviance plot

### Relative influences of features in predicting model

#### Relative Influence

**Table 3.** Relative importance of features.

|  | Relative Influence |
| --- | --- |
| ASA | 59.277 |
| Bloodprodcuts | 20.115 |
| Operativetime | 13.728 |
| Colorectal | 3.714 |
| AGE | 3.165 |
| Major | 0.000 |
| Malignant | 0.000 |
| Gradeofsurgery | 0.000 |

| Relative Influence |  |
| --- | --- |
| Intraophypotension | 0.000 |
| Openlap | 0.000 |
| Smallbowel | 0.000 |
| UPPERGI | 0.000 |
| EMMERGENCY | 0.000 |
| HPB | 0.000 |
| Hernia | 0.000 |

#### Relative Influence Plot

**Figure 3.**
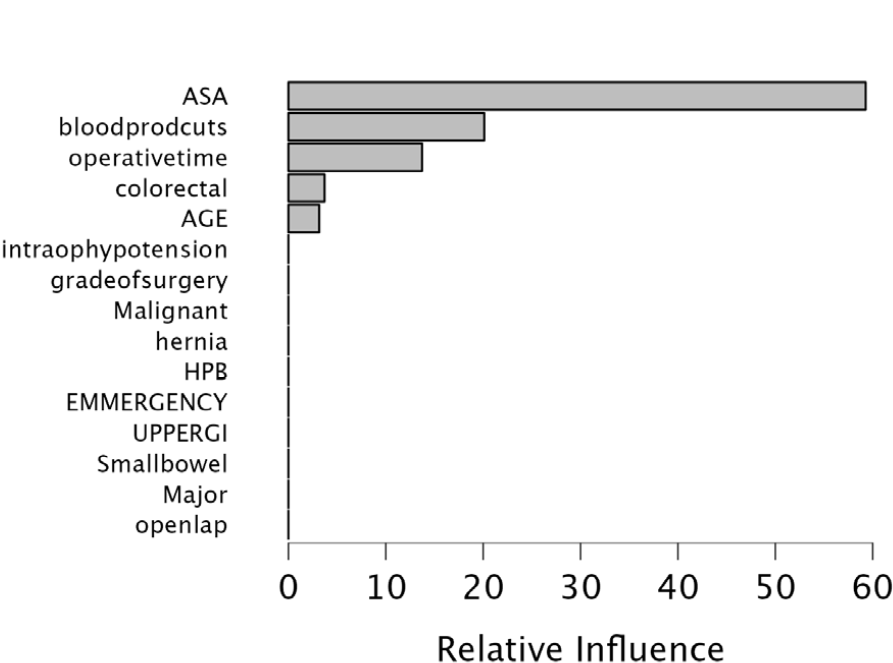
Relative influence of individual features in predicting mortality.

From the relative influence table, it can be seen that ASA grading was the most important feature in predicting mortality (59.27%) followed by blood products (20.11%), operative time (13.72%), colorectal surgery (3.71%) and Age of the patient (3.16%) respectively. It seems the rest of the features almost did not affect 90 days mortality.

### Random forest analysis

We are mentioning in detail the Random Forest analysis algorithm which also showed an overall higher accuracy of 96%, precision of 96.2 and recall of 96.2%. As shown in Table 4, 321 patients’ data were used to train the model. 81 in the validation cohort and 100 in the test cohort.

#### Random Forest Classification

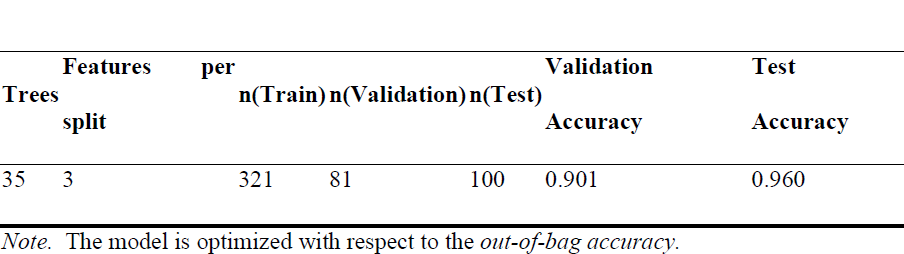

Table 4 Training, validity, and test numbers in random forest classification algorithm,

### Evaluation Metrices

Table 5 shows the evaluation matrices of the model, all other indices are acceptable but it shows high false negative rates in the mortality group again due to the model’s inherent bias towards the group with a high sample size and naturally, as overall mortality was 6%, so the model is biased towards survival, which is one of the limitations of these algorithms, however, to evaluate in such cases Area under ROC curve is taken which shows 0.780 in survival group and 0.754 in mortality group with an average of 0.767. The ROC curve is shown in Figure 4.

**Table 5.**
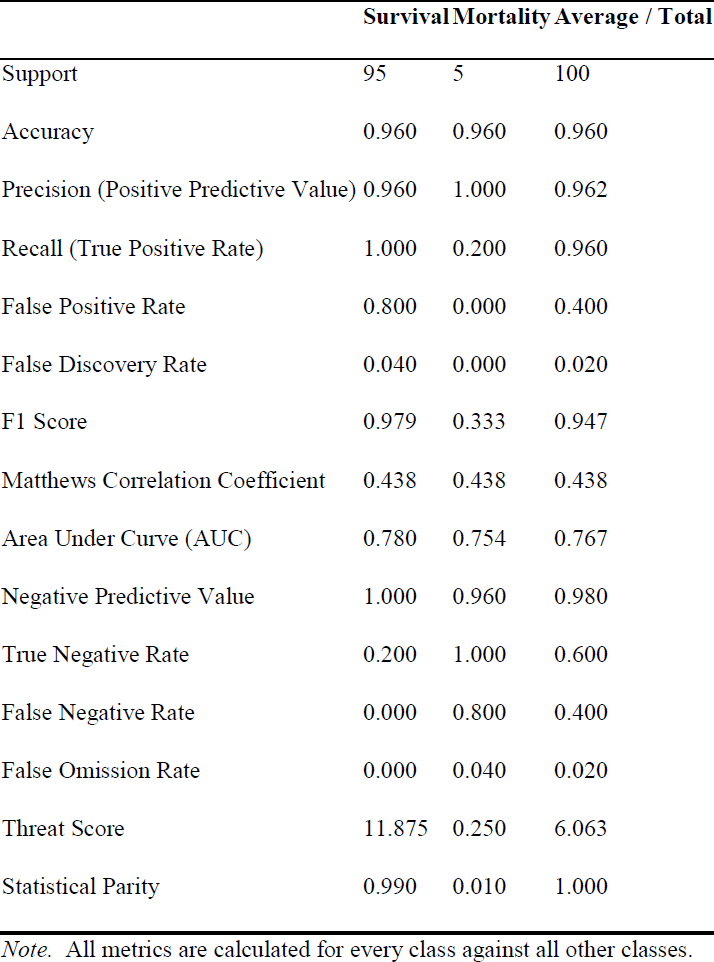
Evaluation Metrics.

#### ROC Curves Plot

**Figure 4.**
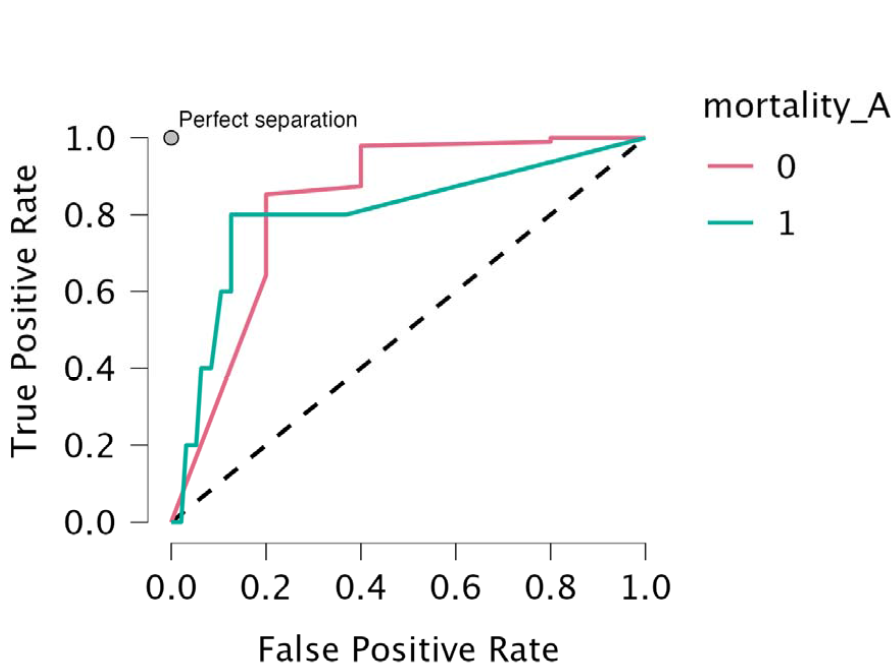
ROC curve red line shows survival group and green mortality group.

#### Out-of-bag Classification Accuracy Plot

Figure 5 shows out of bag accuracy plot which shows as the number of trees increase accuracy is increased and around the number of 35 trees accuracy stabilises, which shows 35 trees taken in model are adequate.

**Figure 5.**
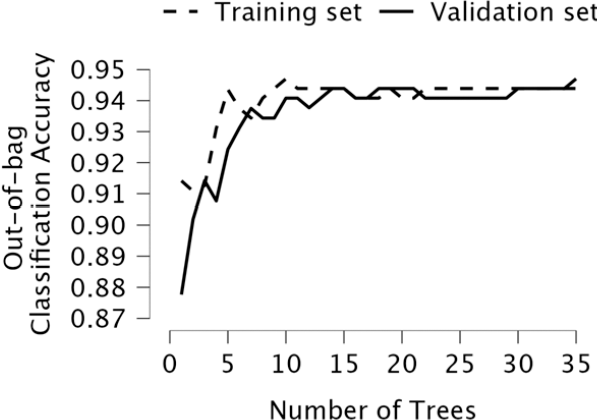
Out of bag accuracy plot Mean Decrease in Accuracy.

#### Mean Decrease in Accuracy

In the random forest algorithm features importance is assessed by mean decrease in accuracy, features which show the highest decrease in accuracy are the most important. In this algorithm also ASA grading was showing the highest importance, followed by open surgery, blood products, age operative time, major surgeries, emergency surgeries, small bowel surgeries, grade of surgeries, HPB surgeries and intraoperative hypotension.

**Figure 6.**
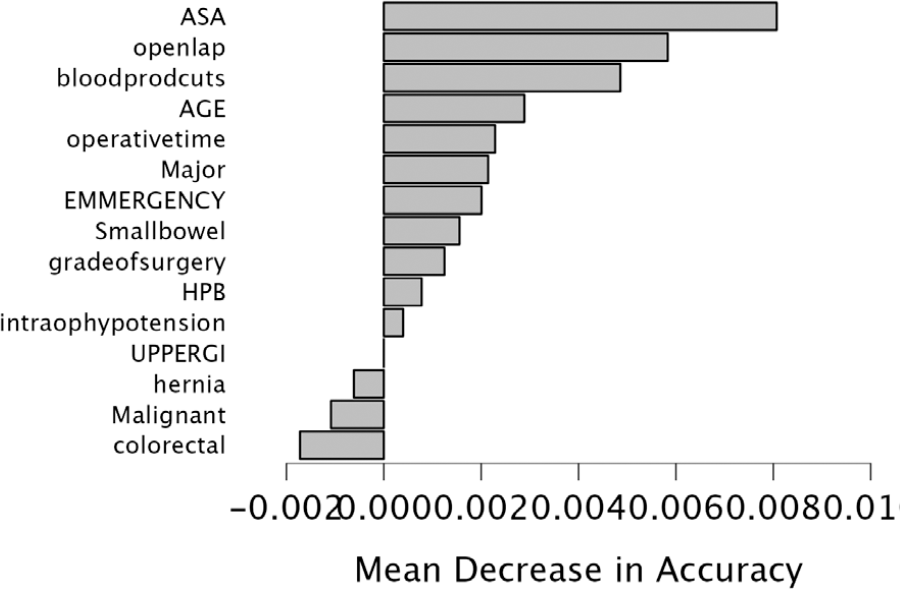
Features importance with mean decrease in accuracy.

### Logistic regression

We compared the accuracy of supervised machine learning models with standardised logistic regression models and their performance diagnostics.

### Performance Diagnostics

Overall accuracy was 95.2% and the area under the curve was 0.703, which showed machine learning models were performing better than standard logistic regression models. Performance metrics.

#### Performance metrics

**Table 6.**
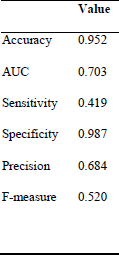
Performance metrics of standard logistic regression model.

#### Performance plots

**Figure 7.**
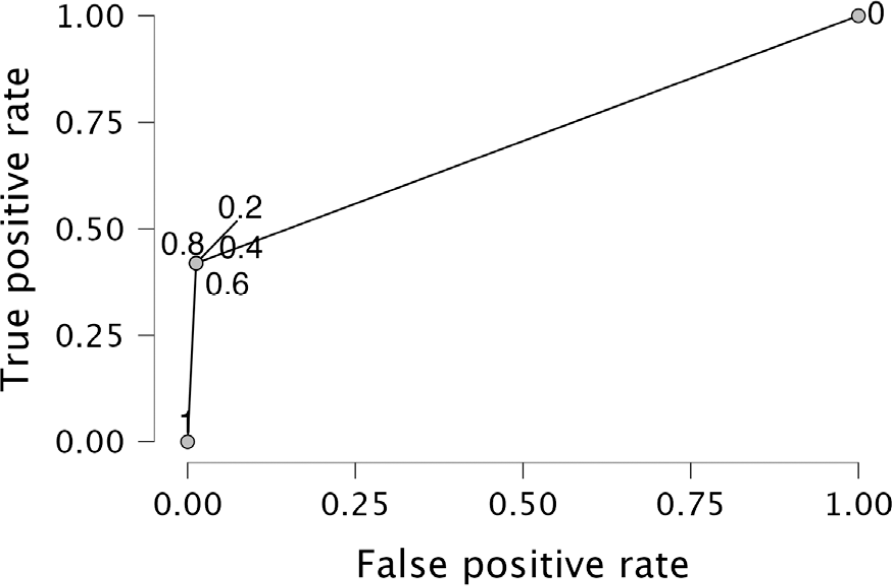
ROC analysis of standard logistic regression model.

## Discussion

Perioperative mortality is one of the most important problems the surgical community must face. Perioperative mortality ranges from 0.1% to as high as 27–30%, depending on the type of surgery [3, 4]. Gastrointestinal and hepatobiliary surgeries are technically demanding procedures and have among the highest perioperative mortality rates. [5,6,7]

Machine learning is a marriage between biostatistics and computer applications. and recently it has gained popularity. There are two kinds of machine learning algorithms supervised and unsupervised. Supervised machine learning algorithms are mainly used for predicting known output or target. [8] There are various scores like (P)POSSUM available to predict postoperative outcomes. [9] However, their accuracies vary according to various centres. One of the benefits of supervised machine learning models is that they can be trained as per our data and can be used to predict outcomes, based on local factors, patients’ profiles etc. and at the same time models created in larger centres can be applied to other centres locally or not locally after checking accuracy.

The primary aim of this study is to create supervised machine learning models based on our data and compare the model with the standard logistic regression method and check for features of importance in predicting the outcomes. We also wanted to evaluate the usefulness of these algorithms in preoperative assessment to predict postoperative outcomes.

As mentioned in the result section we analysed various supervised machine learning algorithms and found that gradient boosting and random forest were predicting the outcomes with the highest accuracy and area under the curve. In gradient boosting American Society of Anaesthesiology, grading was the most important feature, followed by blood products, colorectal surgeries, operative time, and age in decreasing order as shown in Figure 3. In the random forest algorithm, ASA grading was showing the highest importance, followed by open surgery, blood products, age operative time, major surgeries, emergency surgeries, small bowel surgeries, grade of surgeries, HPB surgeries and intraoperative hypotension.

Standard logistic regression showed 95% accuracy compared to 96% accuracy for both gradient boosting and random forest algorithms. It showed an area under the curve of 0.703 compared to 0.857 with gradient boosting and 0.767 for random forest algorithms, which showed supervised machine learning is more or at least as effective as logistic regression.

One of the key limitations of the study is high false negative rates with both the algorithms in the mortality group, but this is the key limitation of classification algorithms when there is a mismatch in class proportion. Overall 90 days mortality was 6.2% so there was a mismatch between survival class and mortality class and hence the finding. As matching the class in mortality analysis was not possible we evaluated the area under the ROC curve, which is another way to evaluate the model in case of a mismatch in class limitation.

In conclusion, supervised machine learning algorithms are highly accurate and precise in predicting post-operative survival and these models can be part of the routine evaluation in predicting post-operative outcomes.

## Data Availability

All data produced in the present study are available upon reasonable request to the authors

